# The predictors of new-onset atrial fibrillation incidence after on-pump coronary artery bypass graft surgery: A prospective study evaluating syntax score

**DOI:** 10.1101/2023.02.21.23286217

**Authors:** Hoda Mombeini, Alireza Ebrahimi, Saeed Yazdankhah, Mohammad Ali Sheikhi, Shahla Majidi, Majid Pakdin

## Abstract

**Background:** Atrial fibrillation (AF) is considered the most common supraventricular arrhythmia in patients undergoing coronary artery bypass graft (CABG). The predictive value of the SYNTAX score for post-CABG new-onset AF incidence has not been clearly evaluated. This study aimed to assess this association in patients undergoing isolated on-pump CABG.

**Method:** This study was done in a single-center, randomized, and observational setting. A total of 133 patients undergoing on-pump isolated CABG who were older than 18 years and had sinus rhythm were enrolled. Demographic variables of patients were recorded, and the SYNTAX score was measured for the participants. The multivariate logistic regression model was applied to identify the predictors of post-CABG new-onset AF.

**Results:** The logistic regression model showed that SYNTAX score of more than 28.25 (p-value= 0.001; OR= 14.25, 95% CI= 2.90_70.11), hypertension (p-value=0.02; OR = 6.59, 95% CI = 1.23_34.57), and calcium channel blocker consumption (p-value=0.02; OR = 8.05, 95% CI = 1.43_45.42) are predictors of new-onset AF after on-pump CABG.

**Conclusion:** This study demonstrated that patients with higher SYNTAX scores in coronary angiography are more likely to develop new-onset AF after isolated on-pump CABG.

## 1. Background

Atrial fibrillation (AF) is the most common arrhythmia occurring after coronary artery bypass graft (CABG) surgery, with an incidence of approximately 27-33% [1-3]. Several risk factors were found to be related to postoperative AF occurrence, such as old age, diabetes mellitus, hypertension, chronic obstructive pulmonary disease, paroxysmal AF history, left ventricular dysfunction, cardiopulmonary bypass time, and cross-clamp time [2]. Patients are more vulnerable to developing AF arrhythmias within the first 4-5 postoperative days, particularly during the first 72 hours after the surgery [4]. Furthermore, post-CABG AF is accompanied by higher complications such as thromboembolism, renal failure, heart failure, mortality, and extended hospital stay, leading to lower quality of life for patients and increased treatment costs [5, 6]. The survival probability was found to be decreased in patients who had suffered from new-onset post-CABG AF [7].

The synergy between percutaneous coronary intervention with taxus and cardiac surgery (SYNTAX) score is based on angiographic imaging that assesses the complexity of coronary lesions and the severity of coronary artery disease (CAD) [8]. This scoring system was first introduced to compare two therapeutic methods of percutaneous coronary intervention with taxus stents versus CABG [6]. SYNTAX is also widely applied to predict the long-term consequences of CAD and decide on appropriate therapeutic interventions [8, 9]. Previous studies mentioned that patients undergoing CABG with higher SYNTAX scores are at greater mortality risk [10]. Although numerous investigations showed that SYNTAX score could be used as a predictive index for several post-CABG complications, such as myocardial infarction and stent thrombosis, little has been done regarding using this score as a predictor of new-onset post-CABG AF. The present study aimed to determine the association of the score with AF incidence after on-pump CABG.

## 2. Methods

### 2.1. Study Design and Population

A total of 133 patients undergoing on-pump isolated CABG surgery at the cardiac surgery ward in Golestan hospital, Ahvaz, Iran, were randomly enrolled in this observational study and were subsequently evaluated for post-CABG AF. Inclusion criteria were defined as individuals older than 18 years old with cardiac sinus rhythm who had CABG. The exclusion criteria included a history of thyroid disease, paroxysmal AF arrhythmia, previous cardiac surgery, percutaneous coronary intervention (PCI) during the past six months, use of amiodarone or digoxin, moderate to severe mitral regurgitation, having implantable cardioverter defibrillator or permanent pacemaker, or left atrial size more than 4 cm.

Patients were observed by continuous electrocardiogram (EKG) monitoring during their stay in the intensive care unit (ICU). After detecting AF during constant monitoring, a 12-lead-EKG was taken to confirm arrhythmia. After being discharged from ICU, patients were also followed in the ward by radial pulse examination at least four times daily in terms of rate and regularity, which was done by expert staff. Should an irregular pulse be observed or patients become symptomatic, a 12-lead-ECG was taken to assess arrhythmia. In this study, AF was described as those that lasted for a minimum of 5 minutes. Ultimately, patients were classified into two groups: patients with AF arrhythmia and those without AF. The patients’ SYNTAX scores compared with each other.

### 2.2. SYNTAX Score Measurement

To calculate the SYNTAX score, we scored each coronary lesion with >50% stenosis in vessels >1.5 mm diameter. Each lesion could involve more than one diseased segment. Other deterministic factors were coronary dominance, total occlusions, trifurcations, bifurcations, aorto-ostial lesions, severe tortuosity, severe calcifications, thrombus, and diffuse diseases.

All angiographic variables were calculated by an expert team consisting of a general cardiologist, an interventional cardiologist, and a cardiac surgeon based on the latest version of SYNTAX score. This version has been designed to improve long-term outcomes prediction following coronary revascularization by subsuming seven clinical variables, including age, peripheral vascular disease, chronic obstructive pulmonary disease, unprotected left main CAD, left ventricular ejection fraction, creatinine clearance, and sex, into the original SYNTAX score [11].

### 2.3. Statistical Analysis

The results were analyzed using Statistical Package for Social Sciences (SPSS), version 22. The relationship between post-CABG AF and different factors was analyzed using Chi-squared test, two-tailed *t*-test, Fisher’s exact test, or Mann-Whitney U test as appropriate. Afterward, the multivariate logistic regression model was performed to identify the predictors of post-CABG AF. The predictive power of SYNTAX score for post-CABG AF was tested by receiver operating characteristic (ROC) curve analysis. P value < 0.05 was considered statistically significant.

### 2.4. Ethical Considerations

The study protocol followed national and international guidelines. Informed consent was obtained from all participants, their anonymity was guaranteed, and the study protocol was approved by the Ethical Committee of Ahvaz Jundishapur University of Medical Sciences (Reg. No. IR.AJUMS.REC.1394.91).

## 3. Results

One hundred thirty-three patients with a mean age of 59 ±8.5 years participated in our study. Most participants were male, and the prevalence rates of diabetes mellitus, hypertension, dyslipidemia, and cigarette smoking were 51.1%, 75.2%, 69.2%, and 25.6%, respectively. The left ventricular ejection fraction (LVEF) ranged from 20% to 60% (Mean ± SD: 47.7 ± 8.8%). The mean number of involved vessels was 2.6 ±0.52, and the SYNTAX score ranged from 7 to 55.5 (mean: 28.66 ±9.95). The left internal thoracic artery was used in 131 (98.5%) patients, while 23 (17.3%) and 80 (60.2%) patients received one and two saphenous vein grafts, respectively. Three and four saphenous vein grafts were used for 25 (18.8%) and 4 (3%), respectively. One remaining patient received no vein graft.

New-onset post-CABG AF incidence in our study was 21.8%. Preoperative variables and medications, as well as perioperative variables, are demonstrated in Tables 1, 2, and 3. During the investigation, there were three cases of mortality which all had developed AF arrhythmia during hospitalization. The cause of two deaths was postoperative acute renal failure. These two cases developed AF on days 2 and 3 of hospitalization. The mortality of the third case, who developed AF on day 7, was due to pneumonia and respiratory failure after prolonged admission in the ICU.

**Table 1.** Pre-operative variables and laboratory data.

|  | Group Non-AF | Group AF | P- value |
| --- | --- | --- | --- |
| | N(%)<br>or Mean ±SD<br>(n=104) | N(%)<br>or Mean ±SD<br>(n=29) | |
| **Age** | 58.7± 8.5 | 60.6± 8.7 | 0.48<sup>1</sup> |
| **Gender (% male)** | 70 (67.3%) | 19 (65.5%) | 0.86<sup>2</sup> |
| **Diabetes Mellitus** | 52 (50.0%) | 16 (55.2%) | 0.62<sup>2</sup> |
| **Hypertension** | 73 (70.2%) | 27 (93.1%) | 0.01<sup>2</sup> |
| **Dyslipidemia** | 71 (68.3%) | 21 (72.4%) | 0.67<sup>2</sup> |
| **Body mass index** | 26.7± 3.4 | 28± 5.3 | 0.24<sup>1</sup> |
| **Cigarette Smoking** | 29 (27.9%) | 5 (17.2%) | 0.25<sup>2</sup> |
| **Stroke history** | 10 (9.6%) | 4 (13.8%) | 0.51<sup>2</sup> |
| **Myocardial infarction history** | 36 (34.6%) | 12 (41.4%) | 0.51<sup>2</sup> |
| **Left main disease** | 12 (11.5%) | 2 (6.9%) | 0.73<sup>2</sup> |
| **Left anterior descending disease** | 104 (100%) | 29 (100%) | >0.99<sup>2</sup> |
| **Left circumflex disease** | 85 (81.7%) | 27 (93.1%) | 0.16<sup>2</sup> |
| **Ramus intermedius artery** | 4 (3.8%) | 1 (3.4%) | 0.92<sup>2</sup> |
| **Right coronary artery disease** | 87 (83.7%) | 25 (86.2%) | 0.74<sup>2</sup> |
| **Number of involved vessels** |  |  | 0.53<sup>1</sup> |
| **One** | 4 (3.8%) | 0 (0.0%) |  |
| **Two** | 28 (26.9%) | 6 (20.7%) |  |
| **Three** | 72 (69.2%) | 23 (79.3%) |  |
| **Left ventricular ejection fraction** | 48.12± 6.76 | 45.17± 9.11 | 0.08<sup>1</sup> |
| **SYNTAX Score** | 27.25± 10.27 | 33.76± 6.66 | 0.012<sup>1</sup> |
| **Creatinine** | 1.01± 0.24 | 1.21± 0.61 | 0.12<sup>1</sup> |
| **Glomerular filtration rate** | 82.93± 24.93 | 76.44± 29.43 | 0.24<sup>1</sup> |
| **Hemoglobin** | 12.3± 1.53 | 12.3± 1.60 | 0.94<sup>1</sup> |
| **Platelet** | 262.38± 77.35 | 299.00± 114.35 | 0.12<sup>1</sup> |

<sup>1</sup> Two-tailed T-test
<sup>2</sup> $\chi^2$

**Table 2.**
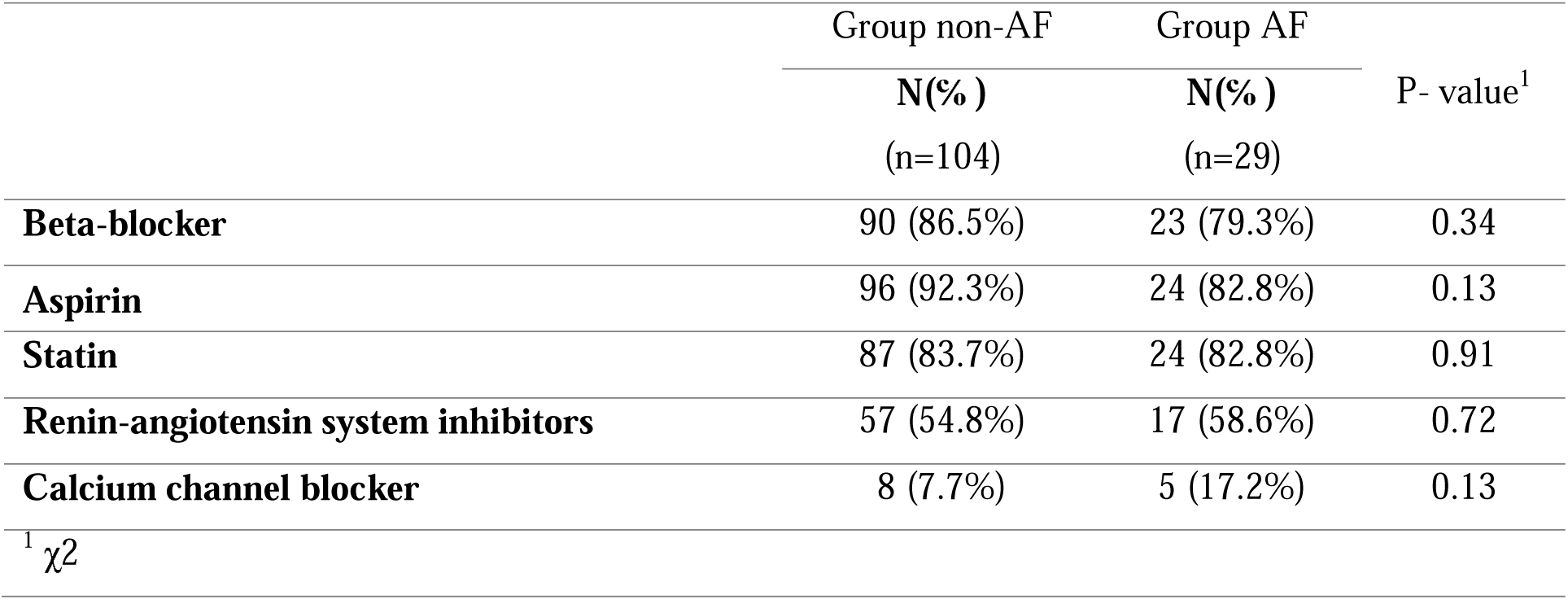
Preoperative medications.

**Table 3.** Peri-operative variables.

|  | Group non-AF | Group AF | P- value |
| --- | --- | --- | --- |
|  | N (%)<br>or mean ±SD<br>(n=104) | N (%)<br>or mean ±SD<br>(n=29) |  |
| **Cardio-pulmonary bypass time** | 74.5±21.93 | 84.00±32.60 | 0.14<sup>1</sup> |
| **Aortic cross-clamp time** | 42.80±11.80 | 43.83±10.15 | 0.51<sup>1</sup> |
| **Use of left internal thoracic artery** | 102 (98.1%) | 29 (100%) | > 0.99<sup>2</sup> |
| **Number of anastomoses** | 2.97±0.74 | 3.31±0.54 | 0.008<sup>1</sup> |

<sup>1</sup> Two-tailed t-test
<sup>2</sup> $\chi^2$

The ROC curve analysis showed good accuracy for the SYNTAX score in predicting post-CABG AF, with the area under the curve being 0.71 [95% CI: (0.69, 0.81) plJ0.001]. The sensitivity and specificity were 89% and 56%, respectively, for SYNTAX scores greater than 28.25 (Figure 1).

**Figure 1.**
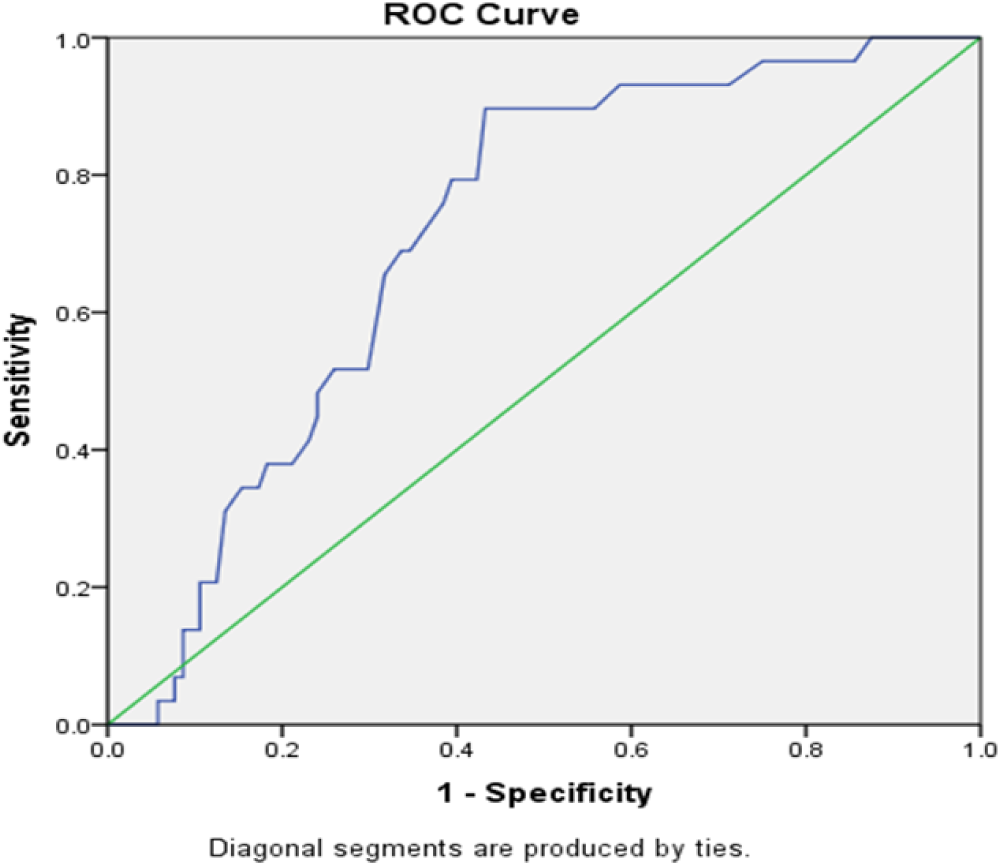
Sensitivity and specificity of SYNTAX Score at Post CABG atrial fibrillation diagnosis.

Multivariate analysis of the risk of new-onset AF incidence was also performed. Ten factors, including aspirin and calcium channel blockers, hypertension, left circumflex (LCX) artery involvement, SYNTAX score, creatinine level, LVEF, platelet count, cardiopulmonary bypass time, and the number of anastomoses were considered in our analysis (Table 4).

**Table 4.** Multivariate analysis of the risk of new-onset AF incidence.

|  | P-value<sup>1</sup> | Odds ratio | 95% CI |
| --- | --- | --- | --- |
| **Aspirin** | 0.210 | 0.336 | 0.62- 1.835 |
| **Calcium channel blocker** | 0.018 | 8.045 | 1.425- 45.416 |
| **Hypertension** | 0.026 | 6.587 | 1.225- 34.568 |
| **Left circumflex artery disease** | 0.677 | 1.472 | 0.238- 9.106 |
| **Syntax score (> 28.25)** | 0.001 | 14.253 | 2.898- 70.111 |
| **Creatinine** | 0.136 | 4.689 | 0.615- 35.737 |
| **Left ventricular ejection fraction** | 0.165 | 0.956 | 0.897- 1.019 |
| **Platelet** | 0.066 | 1.005 | 1.000- 1.011 |
| **Cardiopulmonary bypass time** | 0.330 | 0.988 | 0.963- 1.013 |
| **Number of anastomoses** | 0.090 | 2.471 | 0.869- 7.027 |

<sup>1</sup> Fischer's exact test

The logistic regression model showed that SYNTAX score of more than 28.25 (p-value= 0.001; OR= 14.25, 95% CI= 2.90_70.11), hypertension (p-value=0.02; OR = 6.59, 95% CI = 1.23_34.57), and calcium channel blocker consumption (p-value=0.018; OR = 8.05, 95% CI = 1.43_45.42) were the predictors of AF after on-pump CABG.

## 4. Discussion

AF is considered the most common cardiac arrhythmia and accounts for mechanical heart dysfunction and subsequent complications [12]. The prevalence of AF in the general population has been measured to be 1-2% [12]. Among patients who had CABG, the incidence of AF is much higher, about 25-40% [13], and it has been reported that AF is the leading cause of post-CABG mortality [14]. A range of investigations has examined different prognostic AF indexes in patients undergoing CABG. Consistent with previous findings, our study showed a clinically significant correlation between post-CABG AF and hypertension, use of calcium channel blockers, and high SYNTAX score.

Different scoring systems, including HATCH score, EusoSCORE, and SYNTAX score, have been used to assess the relationship of the obtained score with post-CAB AF incidence in previous studies [12, 13, 15-17]. In the current study, the latter has been used for this purpose. Priorly, SYNTAX score was classified as a reliable predictor of major cardiovascular event rate after revascularization [18-20].

Although the association of SYNTAX score and new-onset post-CABG AF has not been evaluated extensively, few studies have shown the relationship between higher SYNTAX score and new-onset post-CABG AF incidence in patients who underwent on-pump CABG [12, 13, 21]. The present study showed that a higher SYNTAX score could be used as a prognostic factor of new-onset post-CABG AF incidence in patients who underwent on-pump CABG. The cutoff level of SYNTAX score to predict post-CABG AF was 28.25 in our study, which is higher than the determined cutoff level in previous studies [12, 13, 21]. Contrarily, Fukui *et al*. reported that both high SYNTAX score and high Euroscore II were independent predictors of early major complications after CABG; however, according to their investigation, new-onset post-CABG AF was not increased in the group with both high SYNTAX score and high Euroscore II than other groups [22]. Compared to our study, they used a combination of Euroscore II and SYNTAX score, and their study was not designed exclusively to evaluate the correlation between SYNTAX score and new-onset post-CABG AF; as a result, the patients with preoperative AF arrhythmia had not been excluded from the study. Furthermore, the majority of patients in their study had undergone off-pump CABG, and as previously documented, those cases undergoing off-pump CABG are at lower risk of developing AF [23].

Several studies have identified hypertension as a risk factor for post-CABG AF [24, 25]. Consistent with the current body of evidence, our data suggest hypertension as an independent risk factor for post-CABG AF (OR = 6.5). This association may be due to hypertension-related structural changes. A number of changes in the hypertrophied heart, such as fibrosis, may act as a substrate for reentry arrhythmias [26, 27]. Subsequently, AF could be maintained by these focal reentry firings [28].

According to previous studies, non-dihydropyridine calcium channel blockers (CCBs) could significantly reduce the incidence of post-cardiac surgery supraventricular arrhythmias [29]. Additionally, prophylactic diltiazem could reduce new-onset post-CABG AF [30]. In contrast to these findings, a comprehensive evaluation of different pharmacotherapies for the prevention of post-CABG AF reported that CCB therapy could not decrease the odds of new-onset post-CABG AF [31]. In our study, CCB consumers were found to have higher rates of postoperative AF than non-consumers. The following aspects could explain this finding: (1) In our analysis, all patients were on dihydropyridine treatment, not nondihydropyridines. (2) With the paucity of information on postoperative adherence to medications, it may be possible that this result is due to low postoperative continuation of calcium channel blockers and hypertension crisis following calcium channel blocker withdrawal, as the majority of our patients used them for hypertension. There were several limitations in the present study. First, our study population could not be considered representative of all post-CABG patients owing to the fact that our cases were obtained only from one center and are subject to sampling bias. Second, we stated each patient’s cardiac rhythm status based on the preoperative ECG findings relying on the medical charts to determine whether any prior episodes of AF arrhythmia had been noted. As a result, we might have missed potential asymptomatic paroxysmal AF arrhythmia in individuals who showed normal sinus rhythm on the preoperative ECG. Finally, continuous Holter ECG monitoring was not continued after ICU discharge; however, radial pulses and symptoms were assessed in the ward at least four times daily, so there is little likelihood of sustained episodes of AF being missed.

## 5. Conclusion

The results of this study reveal that SYNTAX scorelJ28.25, hypertension, and use of calcium channel blockers are predictors of post-CABG AF. Therefore, it seems sensible to calculate preoperative SYNTAX scores and consider using anti-arrhythmic agents for those with high SYNTAX scores, which may ultimately decrease post-CABG AF occurrence.

## Data Availability

All data produced in the present study are available upon reasonable request to the authors.

## 6. Acknowledgement

The authors appreciate the nurses and physicians at the cardiac surgery ward of Golestan Hospital, Ahvaz, who helped to conduct this study.

